# A Human-Pathogen SEIR-P Model for COVID-19 Outbreak under different Intervention scenarios in Kenya

**DOI:** 10.1101/2020.05.15.20102954

**Authors:** Viona Ojiambo, Mark Kimathi, Samuel Mwalili, Duncan Gathungu, Rachel W. Mbogo

## Abstract

This work describes the mathematical modelling and dynamics of a novel Coronavirus disease 2019 (COVID-19) in Kenya. The mathematical model assumes Human-Human infection as well as Human-Pathogen interaction. Using the SEIR (Susceptible-Exposed-Infected-Recovered) compartmental model with additional component of the pathogen,we simulated the dynamics of COVID-19 outbreak and impact of different control measures. The resulting system of ordinary differential equations (ODEs) are directly solved using a combination of fourth and fifth-order Runge-Kutta methods. Simulation results indicate that non-pharmaceutical measures such as school closure, social distancing and movement restriction emphatically flatten the epidemic peak curve hence leading to a smaller number of overall disease cases.

## Introduction

COVID-19 is an infectious disease that is caused by SARS-CoV-2 which is a newly emergent virus that has attracted alot of scientific research. The first reported case began in mainland China, City of Wuhan, Hubei on 29th December 2019. [1]. Later on, the novel disease began to spread to countries in contact with China resulting to WHO declaring it as a Public Health Emergency of International Concern (PHEIC) on 30 January 2020 [2]. As of 17th April 2020 the number of reported confirmed cases had exceeded two million [3]. The United States of America (USA) was on the lead with infections while Italy followed put. Africa was also affected by imported cases that resulted to the first Kenyan case being reported on the 13th of March 2020 [4].

The three main types of infections of COVID-19 are asymptomatic, pre-asymptomatic and symptomatic. The incubation period for COVID-19 in the pre-assymptomatic infectious group, which is the time between exposure to the virus (becoming infected) and symptom onset, is on average 5-6 days, however it can be up to 14 days. During this period some infected persons can be contagious. In a symptomatic COVID-19 case, the disease manifests itself with signs and symptoms. Asymptomatic transmission refers to transmission of the virus from a person, who does not develop symptoms [5]. The basic reproductive number, defined as the average number of secondary infections produced by a typical case of an infection in a population where everyone is susceptible [6], is affected by the rate of contacts in the host population, the probability of infection being transmitted during contact and the duration of infectiousness. The basic reproductive number for COVID-19 in Kenya ranges from 1.78(95%CI 1.44-2.14) to 3.46 (95%CI 2.81-4.17) [7].

The outbreak is thought to spread through a population via direct contact with infectious individuals [8] or cross transmission through pathogen contamination in the environment. Specifically, the virus is primarily spread between people during close contact,often via small droplets produced by coughing,sneezing, or talking [9] [10] [5] [6] [13]. After breathing out produces these droplets, they usually fall to the ground or on to surfaces rather than remain in the air over long distances [9] [6] [7]. People may also become infected by touching a contaminated surface and then touching their eyes, nose, or mouth. [9] [10]. The virus can survive on surfaces for up to 72 hours [13].

This kind of transmission has had adverse social and economic impact because all segments of the population are significantly affected. The disease is particularly detrimental to vulnerable social groups such as older age-groups, persons with disabilities and people with other underlying diseases. If the pandemic is not properly addressed through policy, the social crisis generated from COVID-19 could also increase inequality, exclusion, discrimination and global unemployment in the medium and long term [11]. Inorder to achieve suitable control measures of COVID-19, an accurate SEIR mathematical prediction model is required. Regrettably, this can only be obtained after the outbreak ends since the prediction trends are largely influenced by each country’s policy and social responsibility [12].

Mathematical models can help us to understand how SARS-CoV-2 could spread across the population and inform control measures that might mitigate future transmission [13]. Non-pharmaceutical interventions based on sustained physical distancing have a strong potential to reduce the magnitude of the epidemic peak of COVID-19 and lead to a smaller number of overall cases. In the absence of a vaccine social distancing has become the most appropriate Non Pharmaceutical Intervention (NPIs) [14]. Lowering and flattening of the epidemic peak is paramount, as this reduces the acute pressure on the health-care system. Premature and sudden lifting of interventions could lead to an earlier secondary peak, which could be flattened by relaxing the interventions gradually [15].

According to the WHO/China Joint Mission Report, 80% of laboratory-confirmed cases in China up to 20 February 2020 have had mild-to moderate disease – including both non-pneumonia and pneumonia cases; whilst 13.8% developed severe disease and 6.1% developed to a critical stage requiring intensive care [16]. It is therefore paramount to come up with an epidemiological model that factors in hospitalization of the mild to moderate cases and severity of the disease.

In a recent study we explored an SEIR-P model that incorporates human behavior during the disease outbreak, such as ignorance of social distancing so as to advise health experts was developed. The model also accounted for individuals with robust immune [17]. The model in this study is different because (i) It does not include individuals with robust immunities because this research has not been fully concluded. (ii) Compartments of hospitalized population *H*(*t*), human population in Intensive Care Unit *U*(*t*) and the fatalities *D*(*t*) have been added to the SEIR-P model so as to account for the aforementioned dynamics [16]. (iii)It investigates the effects of lifting non-Pharmaceutical interventions such as school closure, physical distancing and lockdown on the model. It is envisaged that the dynamics of the results shall inform health experts in their research of efficient and effective ways of flattening the curve.

## Materials and methods

### Model formulation

In this study a human-Pathogen SEIR-P model for COVID-19 outbreak is formulated. The model considers a short-time period thereby ignoring the demographics of births and natural death. The total human population is subdivided into the susceptible human beings *S*(*t*), the exposed human beings *E*(*t*), the asymptomatic infectious population *I_A_*(*t*), the symptomatic infectious population *I_M_* (*t*), the critical infectious population *I_C_*(*t*), the hospitalized population *H*(*t*) in ordinary wards and *U*(*t*) in Intensive Care Unit, the fatalities *D*(*t*) and the recovered population *R*(*t*). The Pathogen vector in the environment is denoted by *P*(*t*).

The model in Figure 1 culminates to a ten dimensional system of ordinary differential equations as follows;

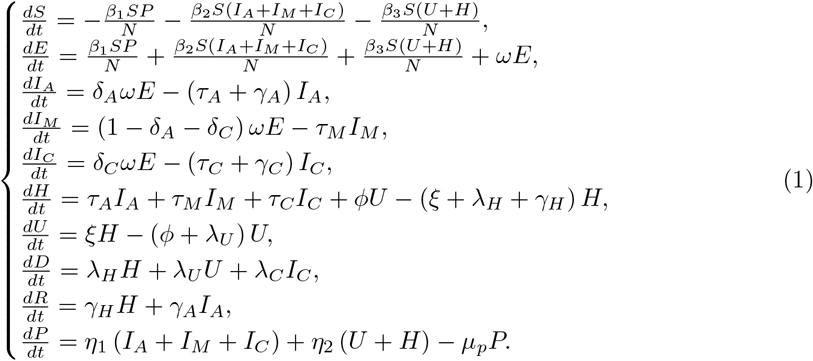

with the initial conditions:

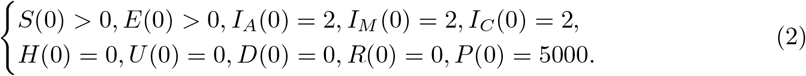

The terms 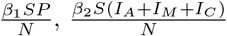 and 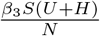 denote the rate at which the susceptible individuals *S*(*t*) gets infected by the pathogens from environment *P*(*t*), infectious humans *I_A_*(*t*), *I_M_*(*t*) and *I_C_*(*t*), and hospitalized humans *U*(*t*) and *H*(*t*) respectively. A quarter of reported cases of COVID-19 are mainly asymptomatic infectious, *I_A_*(*t*) while the larger group is mild infectious, *I_M_*(*t*) with a smaller proportion being critical infectious, *I_C_*(*t*). *I_A_*(*t*) may join the recovery group *R*(*t*) or seek medical intervention from *H*(*t*). *I_M_*(*t*) and *I_C_*(*t*) may go to hospital *H*(*t*), but upon critical diagnosis be referred to the ICU *U*(*t*) which may result to fatalities *D*(*t*) or recovery *R*(*t*). The table of parameters is as shown below. The parameters used in the COVID-19 transmission model are given in the table 1 below.

**Fig 1.**
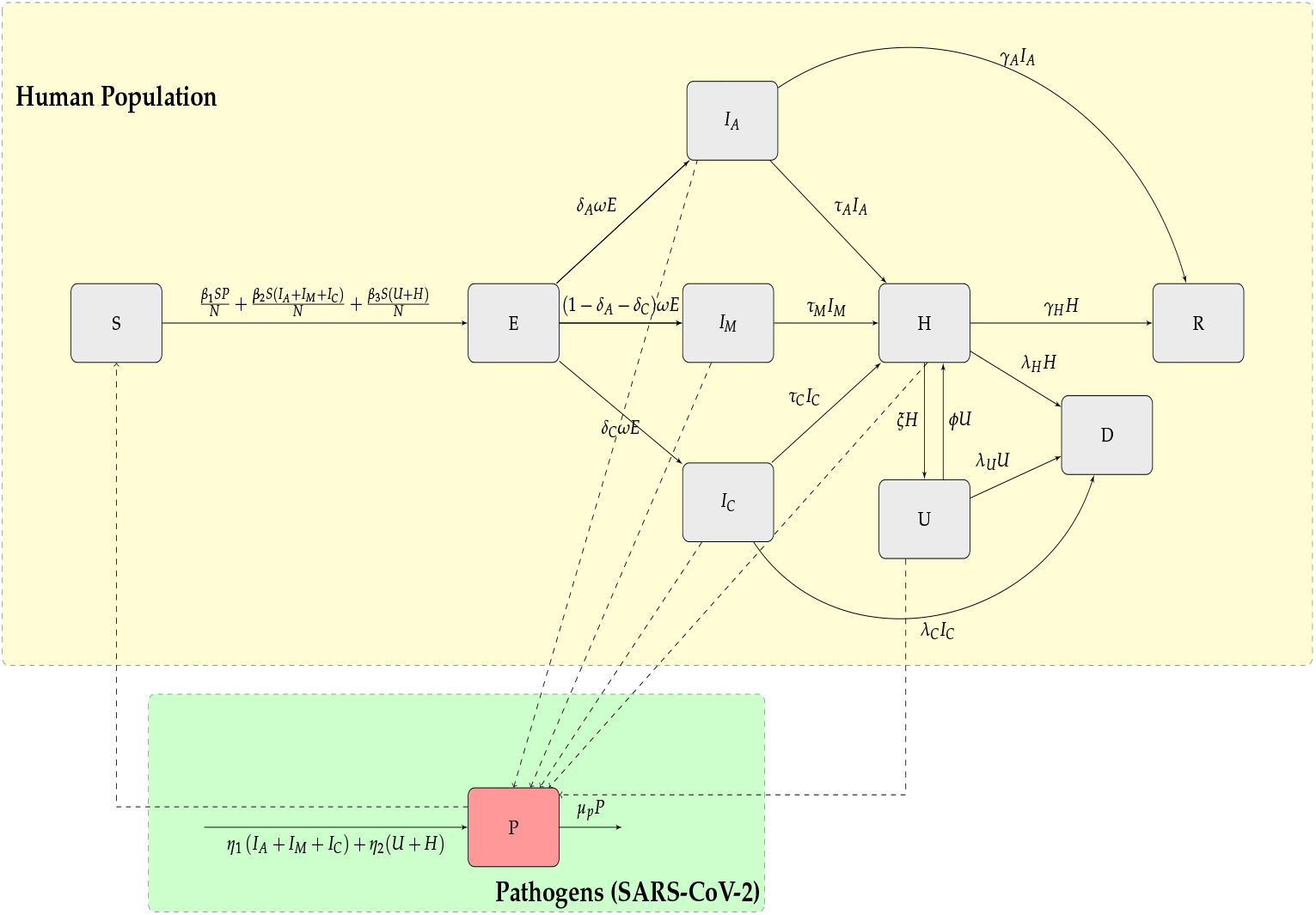
Compartmental model of the SEIR model of transmission of COVID-19

**Table 1.** Description of model parameters.

| Model Parameter | Symbol | Value |
| --- | --- | --- |
| Rate of transmission from $S$ to $E$ due to contact with $P$ | $\beta_1$ | 0.45 |
| Rate of transmission from $S$ to $E$ due to contact with $I_A$, $I_M$ and $I_C$ | $\beta_2$ | 0.75 |
| Rate of transmission from $S$ to $E$ due to contact with $U$ and $H$ | $\beta_3$ | 0.5 |
| Proportion of assymptomatic infectious people | $\delta_A$ | 0.75 |
| Proportion of critical symptomatic infectious people | $\delta_C$ | 0.02 |
| Progression rate from $I_A$ to $H$ | $\tau_A$ | 0.02 |
| Progression rate from $I_M$ to $H$ | $\tau_M$ | 1 |
| Progression rate from $I_C$ to $H$ | $\tau_C$ | 0.99 |
| Progression rate from $E$ to either $I_A$, $I_M$ or $I_C$ | $\omega$ | 0.2 days<sup>-1</sup> |
| Rate of recovery from $I_A$ to $R$ | $\gamma_A$ | 0.98 |
| Rate of recovery from $H$ to $R$ | $\gamma_H$ | 0.97 |
| Progression rate from $H$ to $U$ | $\zeta$ | 0.02 |
| Progression rate from $U$ to $H$ | $\phi$ | 0.2 |
| Progression rate from $U$ to $D$ | $\lambda_U$ | 0.8 |
| Progression rate from $H$ to $D$ | $\lambda_H$ | 0.01 |
| Rate of virus spread to environment by $I_A$, $I_M$ and $I_C$ | $\eta_1$ | 0.55 days<sup>-1</sup> |
| Rate of virus spread to environment by $U$ and $H$ | $\eta_2$ | 0.25 days<sup>-1</sup> |
| Natural death rate of pathogens in the environment | $\mu_P$ | 0.3 days<sup>-1</sup> |

### Disease Free Equilibrium (DFE) and Basic Reproduction Number

In the DFE there are no infections from the humans nor the pathogens such that *S* = *N*. This implies that *P* = *I_A_* = *I_M_* = *I_C_* = *U* = *H* = 0 which further implies that *E* = *D* = *R* = 0.

*I_A_*The Basic Reproduction Number *R*_0_, is defined as the average number of secondary infections produced by a typical case of an infection in a population where everyone is susceptible [6]. This number is affected by the rate of contacts in the host population, the probability of infection being transmitted during contact and the duration of infectiousness. The next generation matrix method is used to obtain the *R*_0_.

Let *x* = (*E*, *I_A_*, *I_M_*, *I_C_*, *H*, *U*, *P*)*^T^* then the model can be written as 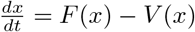, where

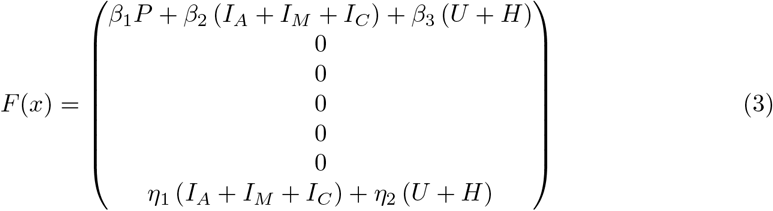

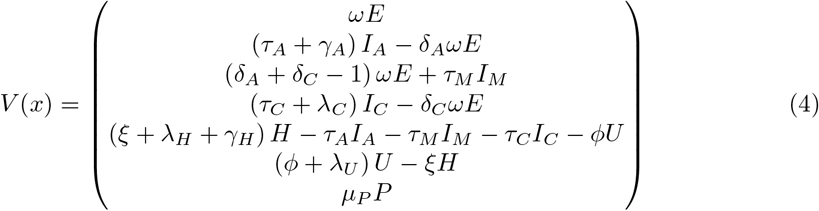

Evaluating the derivatives of F and V at the Disease Free Equilibrium yields matrix **F** and **V** as follows;

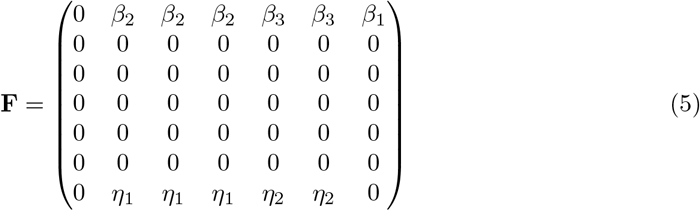

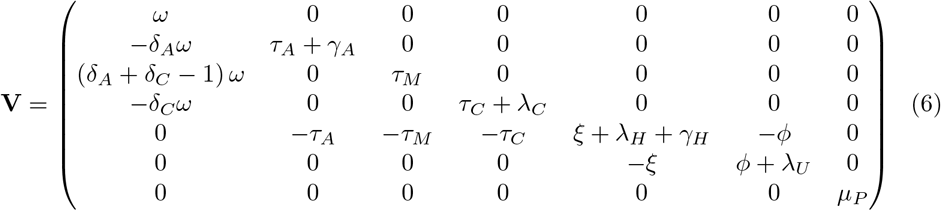

Substituting, (*ξ* + λ*_H_* + *γ_H_*) (*ϕ* + λ*_U_*) = *C*_5_*C*_6_ − *ξϕ = C*_1_, *λ_C_* + *τ_C_ = C*_2_*,τ_A_ + γ_A_ = C*_3_, (*ϕ + λ_U_*) − *ξϕ* = *C*_6_ − *ξϕ* = *C*_4_, *ξ + λ_H_ + γ_H_ = C*_5_ and *ϕ +* λ*_U_* = *C*_6_ in matrix **V** yields

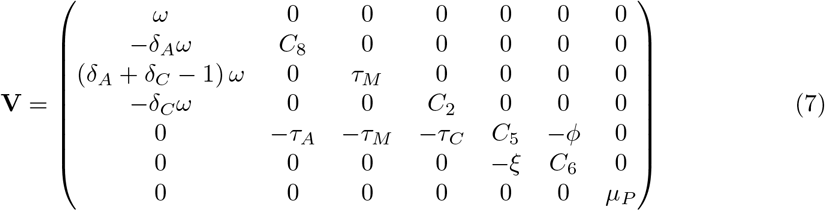

The inverse of matrix **V** is determined as

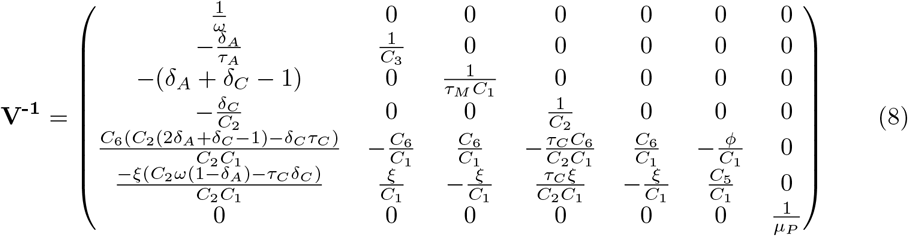

The product **FV^-1^** results to;

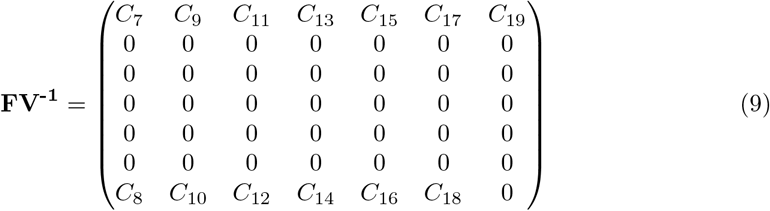

where

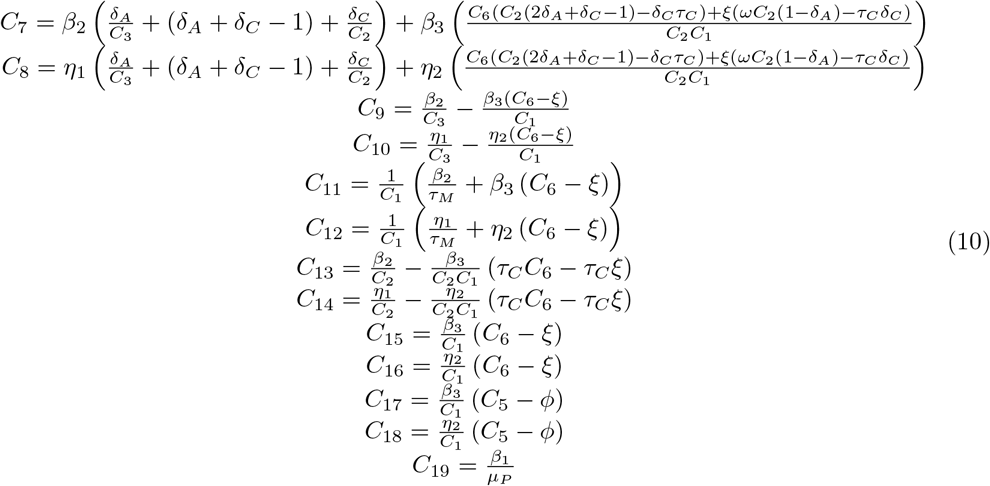

*R*_0_ is the spectral radius of the product **FV^-1^** that results to;

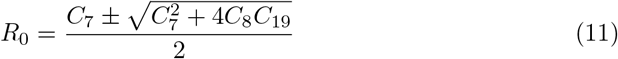

Here *R*_0_ is the sum of three terms representing the average new infections contributed by each of the three infectious classes; pathogens, infectious groups (mild, assymptomatic and critical) and hospitalized groups denoted by *β*_1_, *β*_2_, and *β*_3_ respectively.

## Results and Discussion

A cure or vaccine for COVID-19 is yet to be discovered. Therefore, the interventions being carried out are non-pharmaceutical and include measures such as contact tracing, quarantine, social distancing, wearing of face masks, and hand washing. Mass testing and effective contact tracing has been a great challenge in low-income countries. Therefore, these countries have adopted social distancing measures such as closure of schools, reducing sizes of gatherings, minimizing contact in workplaces, and imposing curfews. The main aim of such interventions is to flatten the curve i.e. prevent infection from spreading fast and bring down the number of infected cases. Curve flattening is necessary because it avoids overwhelming the health systems and gives them time to develop treatment mechanisms.

A key factor in slowing infection spread is reduction of the basic reproduction number *R*_0_, which is expressed in terms of the rates of new infections *β*_1_, *β*_2_, and *β*_3_. Thus, we implement the non-pharmaceutical interventions (NPIs) by multiplying the rates of infections by a 20% and 30% factor. The interpretation is that the NPIs yield an overall mitigation of the epidemic spread by 20% and 30% respectively. An attack rate of 67% was assumed. This led to considering total population *N* as the product of attack rate and the total population of Kenya, which is approximately 5.02 × 10^7^. An identification rate *p* of the new infectious cases was determined as the ratio of confirmed cases over the total number of tested individuals. At the time of this study *p* was 0.02, while the parameter values were taken as shown in Table 1 above. The simulation was initialized at date 13-3-2020 and run for 365 days with the initial values *I_A_*(0) = 2, *I_M_*(0) = 2, *I_C_*(0) = 2, and *P*(0) = 5,000, while the other variables (except *S*(0)) were initially set to 0.

The results of the infectious (*I_A_* + *I_M_* + *I_C_*) and hospitalized (*H* + *U*) cases in Kenya are shown in Figure 2, and epidemic peak values and dates are depicted in Table 2. In unmitigated situation, the peak arrives rather quickly, in about 4 months probably due to the asymptomatics significantly contributing to the epidemic spread. We note that the Kenyan population is largely comprised of youth, who are majorly presumed to be an asymptomatic population. Our model predicts that if the overall control measures achieves 20% effectiveness in slowing the spread of COVID-19, then a delay of slightly 2 months is realized, which allows the health care systems time of gathering resources to manage the epidemic. Consequently, the reported infectious and hospitalized cases are reduced by more than 50%. However, there is a likelihood of exceeding the health capacities. A 30% effectiveness of the applied NPIs substantially bring the epidemic further down, and as noted in Figure 3 the simulated cumulative infectious cases largely coincides with the data of COVID-19 epidemic in Kenya. Moreover, this particular mitigation effort reduce the cumulative infectious cases from just under 700,000 down to around 450,000 cases by date 13-Mar-2021. A similar reduction is also seen in the cumulative number of hospitalized cases. Moreover, the cumulative deaths attains a more than 50% reduction, from about 6,000 cases in unmitigated situation to around 3,500 cases in the 30% mitigation scenario. Majority of deaths emanate from those already in intensive care unit (ICU) *U*, which is presumably overwhelmed by the many critically ill patients arising from *H* and *I_C_* populations. The inability to access the limited ICU resources results to additional deaths from *H* and *I_C_* populations.

**Fig 2.**
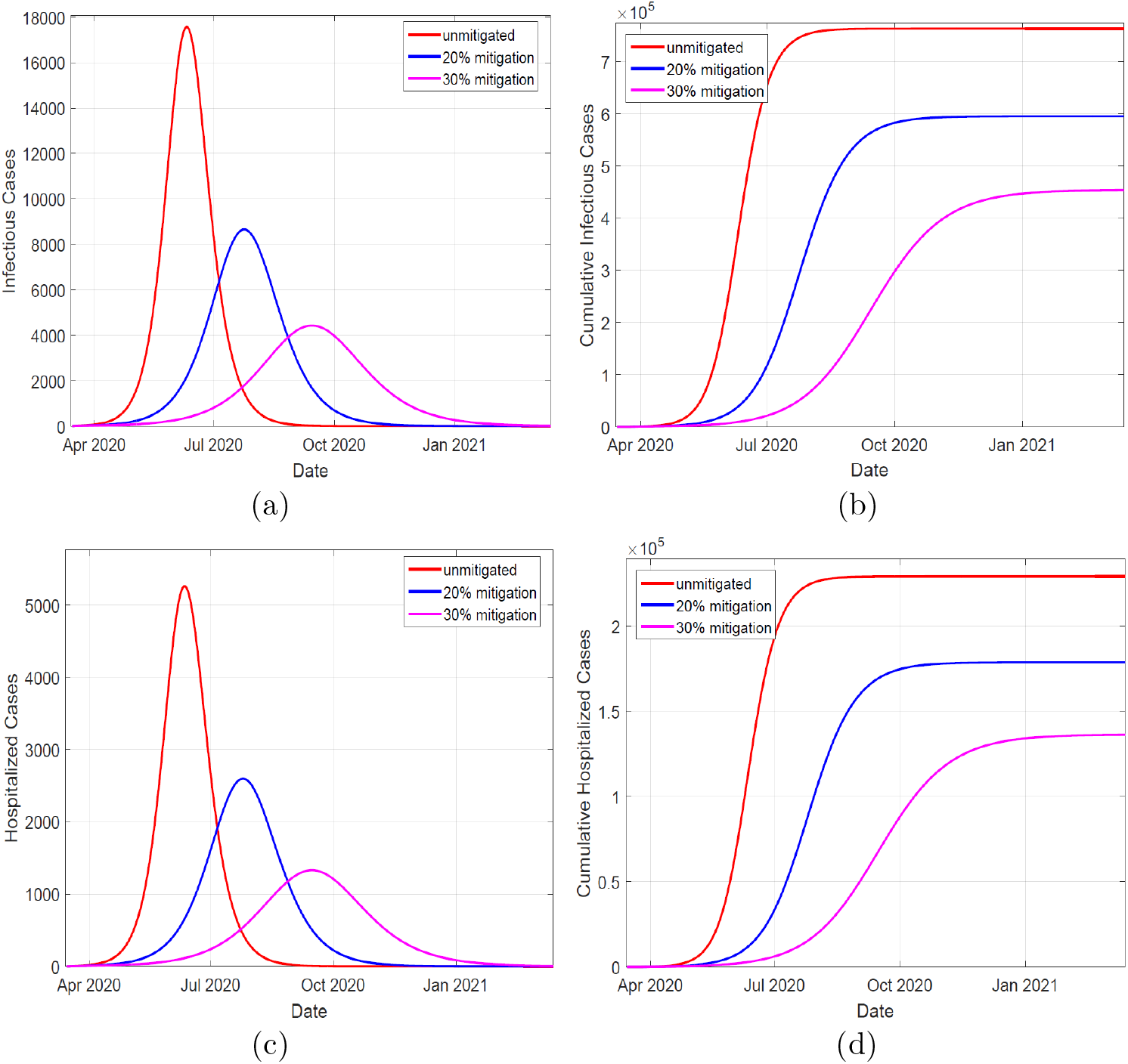
Left panels depicts simulation of infectious and hospitalized cases, in unmitigated and mitigated situations. The implementation of NPIs results to a delay and significant reduction in the cases, as shown by the varying peaks. Right panels show the simulated cumulative cases for both infectious and hospitalized individuals. The infectious cases comprise of individuals in compartments *I_A_*, *I_M_*, and *I_C_* cases.The hospitalized cases comprise of individuals in compartments *H* and *U*.

**Table 2.**
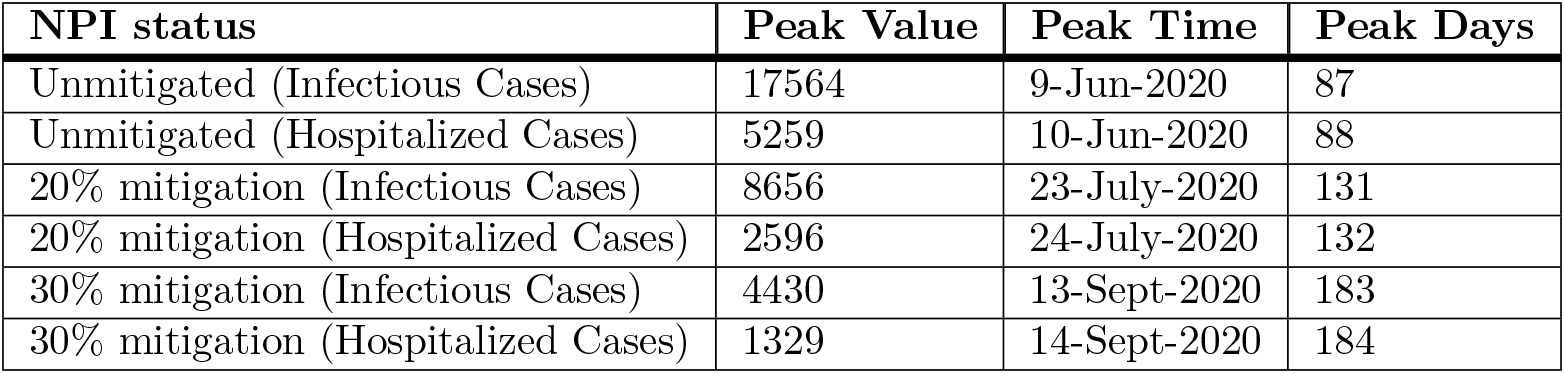
Simulated peaks of infectious and hospitalized cases, with the associated peak dates.

**Fig 3.**
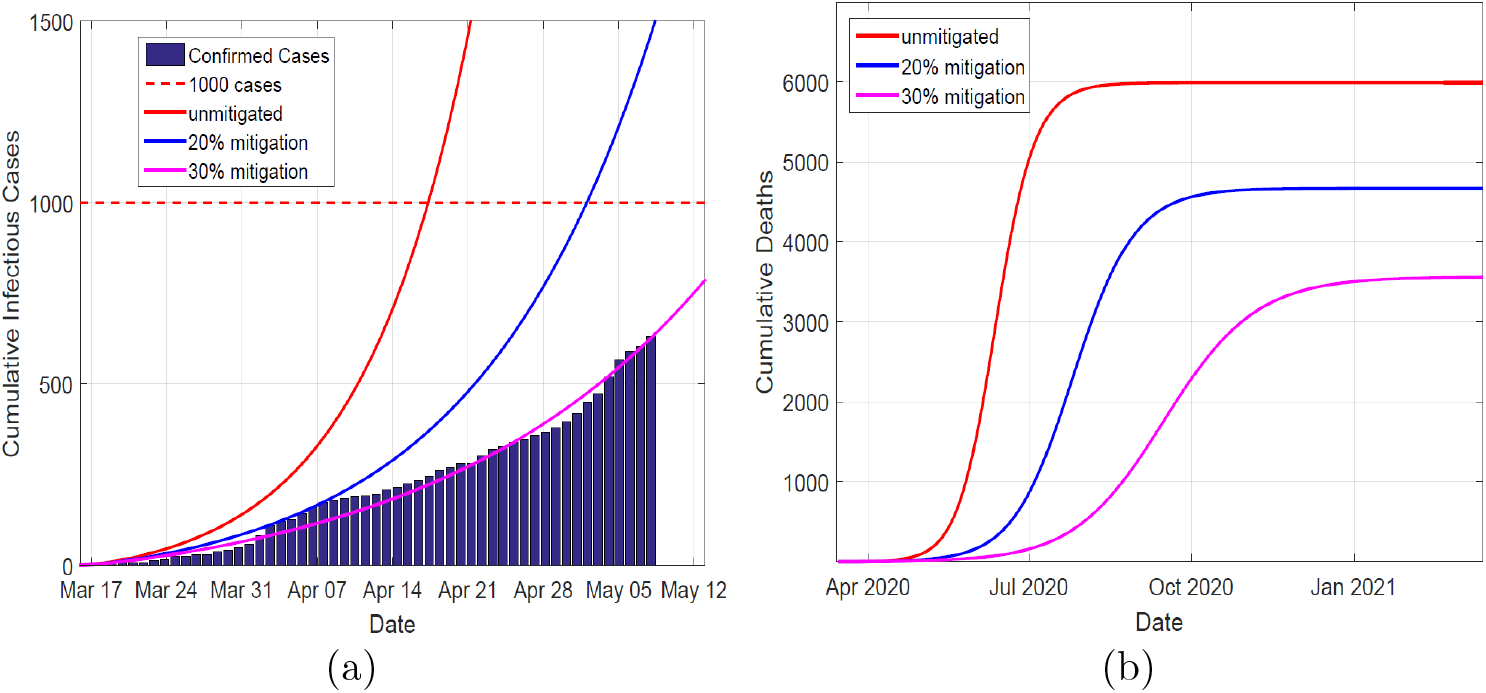
Left panel depicts the data of the confirmed cases in Kenya against the simulated infectious cases for the first 60 days, in unmitigated and mitigated situations. The simulated results of a 30% NPIs effectiveness coincides with the data. Right panel show the cumulative deaths, which decline significantly as the NPIs take effect.

The control measures were put in place for the entire simulation time, so there was no rebound of the infections. A strong one-time social distancing measure would not sufficiently prevent overwhelming of the health care capacities, since it keeps a significant number of the population in susceptible compartment such that a rebound in infections, after the measure has been lifted, will lead to an epidemic wave that surpasses the capacity. Moreover, previous studies have shown that intermittent application of the NPIs maintains the infections within the health care capacities, but prolongs the total duration of the epidemic.

The table depicts the predicted peak values of (daily)reported cases and corresponding times. The cases are placed in two categories, namely: infectious (*I_A_* + *I_M_* + *I_C_*) and hospitalized (*H* + *U*)

## Conclusion

An SEIR-P compartmental model has been used to study the spread of COVID-19. The model accounts for both human to human and human-environment interactions. The model simulations underscores the fact that NPIs such as school closure, social distancing and movement restriction emphatically delay or flatten the epidemic peak.

## Data Availability

The data used to support findings of this study is available from the corresponding author upon request.

## Acknowledgements

The authors appreciate ample time given by their respective universities towards this manuscript.

